# Prevalence and associated factors of diarrhea among under-five children in Debre Berhan City, internally displaced people’s centers, Ethiopia, 2025: A cross-sectional study

**DOI:** 10.64898/2026.06.19.26356028

**Authors:** Asaye Worku Agegn, Eleni Dagnew, Abebe Nigussie, Tewodros Mulugeta, Hibist Kinfe, Tsegaamlak Kumelachew

**Author notes:** Corresponding Author: Asaye Worku Agegn.

## Abstract

**Background:** Diarrheal disease remains a major public health concern among under-five children, particularly in the displaced population, where sanitation, water supply, and health care access are limited. Although it is a 3^rd^ leading cause of under-five morbidity and mortality, limited data exist on the prevalence and determinants of diarrhea in IDP centers.

**Objective:** To assess the prevalence and associated factors of diarrhea among under-five children in Debre Birhan City Internally Displaced People’s centers in 2025.

**Methods:** A cross-sectional study was conducted from December 16--30, 2025, in Debre Berhan citys internally displaced people centers. A total of 355 mothers/caregivers were selected using systematic random sampling. Data were collected through face-to-face interviews using structured, pre-tested questionnaires via the Kobo Collect application. The data were analyzed by using SPSS version 26. Bivariable and multivariable logistic regression were fitted to identify factors associated with the outcome variable. An adjusted odds ratio with its 95% CI was used to determine the strength of association, and variables with a p-value of <0.05 were considered significant.

**Result:** The prevalence of diarrhea among children under five years in Debre Berehan city internally displaced peoples center was 32.4% (95% CI: 28-37). Three factors, unable to read and write educational status of mothers (AOR =3.1; CI: 1.27-7.6), open dumping waste disposal method (AOR=2.63, 95%CI: 1.27-5.5), unvaccination (AOR=3.9, 95%CI: 1.4-10.85) and partial vaccination (AOR= 2.22, 95%CI: 1.25-3.95) were significantly associated with the outcome variable diarrhea.

**Conclusion:** The prevalence of diarrhea among children under five in the IDP center of Debre Berhan city was high. Management strategies to improve maternal health literacy, promote proper waste management and sanitation, and enhance outreach immunization services are crucial for reducing the burden of diarrhea.

## INTRODUCTION

Globally, 83.4 million internally displaced persons (IDPs) were reported as a result of violence, conflict, and disasters at the end of 2024, where 46% of them were residing in sub-Saharan Africa(1). In Ethiopia, there have been 3,135,000 IDPs due to armed conflicts and disasters in the same year (1). IDPs live in extremely unhealthy conditions due to poor sanitation of water, overcrowded housing conditions, unhygienic habits, malnutrition, and unsafe food management (2). All of these factors increase the risk of developing infectious diseases such as diarrhea (3).

According to the WHO, diarrhea in children is defined as a passage of three or more loose or liquid stools per day, or more frequently than is typical for the individual (4). Diarrheal Disease (DD) is caused by a variety of bacteria, viruses, and parasites, leading to dehydration, malnutrition, and susceptibility to other infections or death within a short period of time (5,6). It causes 1.7 billion cases of diarrhea as a leading cause of under-five death, killing about 443832 children every year (7). It has become a worldwide major public health concern with a high rate of morbidity and mortality, particularly in developing countries such as Sub-Saharan Africa (SSA), including Ethiopia (3,8).

Eventhough, numerious interventions such as clean water access, sanitation facilities, vaccination campaigns, have been implemented to curb the prevalence worldwide, the burden remains disproportionately and unacceptably high in SSA (7). A systematic review and meta-analysis reports in Africa shows that the prevalence of diarrhea among children under-five years was 23.59% in community-based studies and 28% in IDP centers (3,9).

Few studies have been done in the past in developing countries about the prevalence and associated factors of diarrhea among children under-five years in IDP centers, such as 51% in Hargeisa, Somaliland (8), 35% Kahartoum, Sudan (10), and 52.3% Mekele, Ethiopia (11). Various factors such as age, duration of breastfeeding, source of water, handwashing practice, water supply, waste disposal practice, caregivers’ educational status, occupation, were independent predictors of diarrhea occurrence in Ethiopia (2,5,6,12–17).

The prevalence of diarrhea among children under five years in Ethiopia from various community-based studies range from 11-27.3% (5,6,12–17). However, little information is known across the country about the magnitude in IDP centers and furthermore, no study has been conducted in the study area. Therefore, this study aims to assess the prevalence and associated factors of diarrhoea among under-five children in Debre Berhan town IDP centers, North East Ethiopia.

## METHOD AND MATERIALS

### Study area and period

The study was conducted in Debre Berhan IDP centers in Debre Berhan city, North Shewa Amhara Region, Ethiopia. Debre Berhan city is the capital of North Shewa zone and is located 130 km from Addis Ababa. Geographically, it lies between latitude 9°41′ N and longitude 39°32′ E and lies at an altitude of 3000 m above sea level. In Debre Berhan city administration, there are 3269 under five children living in three IDP centers (as of December 2025), namely Bakello, Woynshet, and China Camp. The study was conducted from December 16-30, 2025.

### Study design

A descriptive cross-sectional study design was employed.

#### Population

The source population were all children under-five years old who were living in Debre Berhan town IDP center, and the study population was selected under-five children who were living in Debre Berhan town IDP centers. The study unit consists of each randomly selected under five children who lived in Debre Berhan IDP centers during the data collection period.

### Illegibility Criteria

The inclusion criteria were all under-five children with available caregivers who had lived in the IDP household for at least 2 weeks prior to data collection. Whereas critically sick caregivers and those with hearing abnormalities were excluded from the study.

### Sample size determination and sampling technique

#### Sample size determination

In order to determine the study population sample size, the study utilized the single population proportion formula.

n1 = (Zα/2)^2^ (pxq)/d ^2^

✓ P = the proportion of children who have diarrhea, p=0.52 (52.3 %) (11).
✓ Zα/2 =the critical value at the 95% confidence interval level of certainty (1.96)
✓ d = the margin of error between the sample and population, 5% or 0.05 Thus, n_1_ = (1.96)^2^ (0.52×0.48)/0.05^2^ = 383

As this source population was 3,269 less than 10,000, a population correction formula was applied to determine the adjusted sample size.

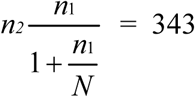

By adding 10% non-respondent rate, which is 34, the final sample size will be =377

### Sampling technique and procedure

The systematic random sampling technique was used to select caregiver/child pairs of participants. K value (the interval between consecutive participants) was calculated by 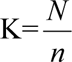 = 9. Where K =interval, N is the total population, and n is the final sample size. Finally, individual study participants were selected using a systematic random sampling technique from the compiled list of all eligible under-five children registered in each IDP center, in which every 9th child was selected. The first number to start (number two) was determined by a lottery method from one to nine. Proportional allocation of the final sample size (n=377) was performed for each camp based on its respective population of under-five children

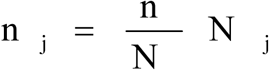

- nj is the sample size of the jth stratum

- Nj is the population size of the jth stratum

- n = n1 + n2 + …+ nk is the total sample size

- N = N1 + N2 + …+ N k is the total population siz

**Figure 1:**
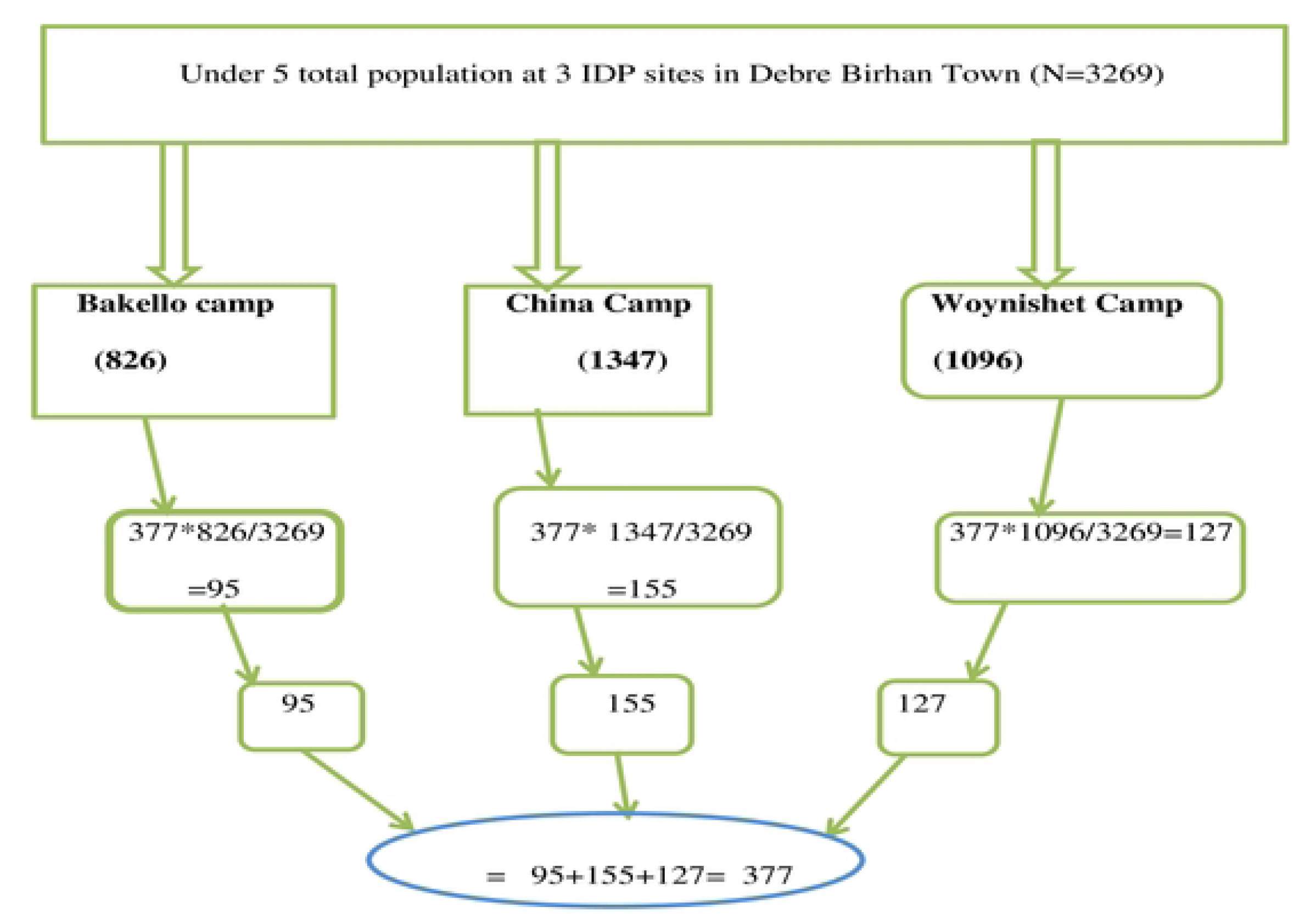
Diagramatic presentation of sampling procedure for under-five children in Debre Berhan IDP center, 2025.

### Study variables

#### Dependent variable

➢ Prevalence of diarrhea

#### Independent variable

A) **Sociodemographic factors:** Age and sex of both child and caregiver, education status of the care giver/ mother, occupational status of the care giver/ mother, source of income and size of the family.
B) **Behavioral factors:** Drinking water separation, hand washing time, handwashing method, method of household waste disposal practice, knowledge of prevention of diarrhea, availability of hand washing facility, method of diarrhea prevention, feeding practices, rota vaccination, breastfeeding experience.
C) **Environmental factor:** Water availability, source of water supply, method of water storage, source of drinking water, latrine utilization**, t**ype of latrine and frequency of soap hand washing.
D) **Accessibility of health facility and child care:** Distance of health facility, visit health care facility, place of care, duration of illness, vitamin A and deworming supplementation, zinc Supplementation, Duration and color of diarrhea, level of dehydration and co-morbidity.
E) **Nutritional factors:** Wasting, stunting, underweight and Mid-upper arm circumference.

### Operational definition

**IDPs:** persons or groups of persons who have been forced or obliged to flee or to leave their homes or places of habitual residence, in particular as a result of or in order to avoid the effects of armed conflict, situations of generalized violence, violations of human rights, natural or human-made disasters, and who have not crossed an internationally recognized State border (11).

➢ Diarrhea: Passage of loose or watery stool three or more times during a 24-hour period (4).

### Data Collection Tool and Procedure

Data were collected by using a pre-tested structured questionnaire (Data collection tool) developed in the Kobo Collect application, which was adapted by reviewing previous studies from the literature. The tool consists of sociodemographic factors, behavioral factors, environmental factors, health facility, and clinical and nutritional factors.

Data collection was conducted from under-five children registration book at each IDP site. Data were collected via face-to-face interviews from mothers or primary caregivers of children under-five by trained one master’s and three degree pediatric nurses.

### Data Quality Control

To ensure data quality, the questionnaire was first written in English and then translated into Amharic, and the translated Amharic version was rewritten in English to ensure consistency and acuracy of data. Prior to data collection, one day training was given to the supervisor and data collectors about the purpose of data collection, the content of the questionnaire, the method of interveiw and ethical issues related to the study participants. The questionnaire was pretested on 5% of the sample size before the actual data collection to ensure for clarity, consistency, and validity out of the study area in Debre Birhan city, Selamchora kebele. Daily supervision and onsite checking of completed questionnaires were carried out by the supervisor and principal investigator.

### Data Analysis and Processing

Prior to data entry, all questionnaires were checked for completeness. Cleaned and coded data were entered into the Kobocollector software and then exported to the Statistical Package for Social Sciences (SPSS) version 26 for analysis. Descriptive characteristics were described in terms of central tendency (mean, median) and dispersion value for continuous data. While Categorical data were described by frequency distribution and presented in the table, text, and figure.

To identify factors associated with diarrhea, bivariable logistic regression was first performed for each independent variable. Variables with a p-value of less than 0.25 in the bivariable analysis were included in the subsequent multivariable logistic regression model. In the final multivariable model, an adjusted odds ratio (AOR) with a p-value of less than 0.05 and its 95% confidence interval (CI) was considered statistically significant.

### Ethical consideration

The Ethical approval was obtained from Debre Berhan University, Asrat Woldeyes Health Sciences Campus, Inistitutional Review Board with Ref number: IRB 01/40/2018. The letter of cooperation were issued to Debre Berhan city administration health office and IDP centers ensuring the approval and neccessary facilitation for smooth undertaking of the study. Participant recruitment was conducted from December 16-30 2025. Written informed consent was obtained from mothers or caregivers after informing them the purpose of the study and their full right to withdraw at any time during data collection. Anonymity and confidentiality of information were guaranteed. Participation was voluntarily and caregivers were informed of their right to withdraw from the study at any time without penality and loss of service. The study was conducted in accordance with declaration of Helsinki.

## Result

### Sociodemographic characteristics of study participants

Among 377 caregivers-child respondents, 355 were included in the study with a response rate of 94%. The majority of caregivers 316 (89%), were females, and more than half 189 (53.2%) of them were below the age of 35 years. The educational background of the mothers or caregivers indicates 105 (29.6%) were unable to read and write, 107 (30.1%) only read and write, 85 (23.9%) had primary education, and 58 (16.3%) of them had attended secondary education and above. Nearly two-thirds 229 (64.5%) of households were living with 4-5 persons per family (Table 1). The mean age of children was 27.47 months. The age category shows about one forth 87 (24.5%) were between 24 and 35 month, and the lowest 27 (7.6%) age category were below six month (Figure 2).

**Table 1:** Sociodemographic characteristics of study participants in Debre Berehan IDP center, (N= 355)

| Variable | Category | Frequency | percent |
| --- | --- | --- | --- |
| Care givers gender | Male | 39 | 11% |
|  | Female | 316 | 89% |
| Care givers age | <35 | 189 | 53.2% |
|  | 35-45 | 120 | 33.8% |
|  | >45 | 46 | 13% |
| Gender of the child | Male | 158 | 44.5% |
|  | Female | 166 | 46.8% |
| Level of education of the mother or caregiver | Unable to read and write | 105 | 29.6% |
|  | Can read and write | 107 | 30.1% |
|  | Primary | 85 | 23.9% |
|  | Secondary | 58 | 16.3% |
|  | Daily laborer | 37 | 10.4% |
| Current occupation of mother/ caregiver | Student | 17 | 4.8% |
|  | House wife | 190 | 53.5% |
|  | I don't have | 111 | 31.3% |
| Family current main source of income | Agriculture | 6 | 1.7% |
|  | Own business | 26 | 7.3% |
|  | From aid | 323 | 91% |
| Family size | 1-3 per house | 54 | 15.2% |
|  | 4-5 per house | 229 | 64.5 % |
|  | Above 6 per house | 72 | 20.3% |

**Figure 2.**
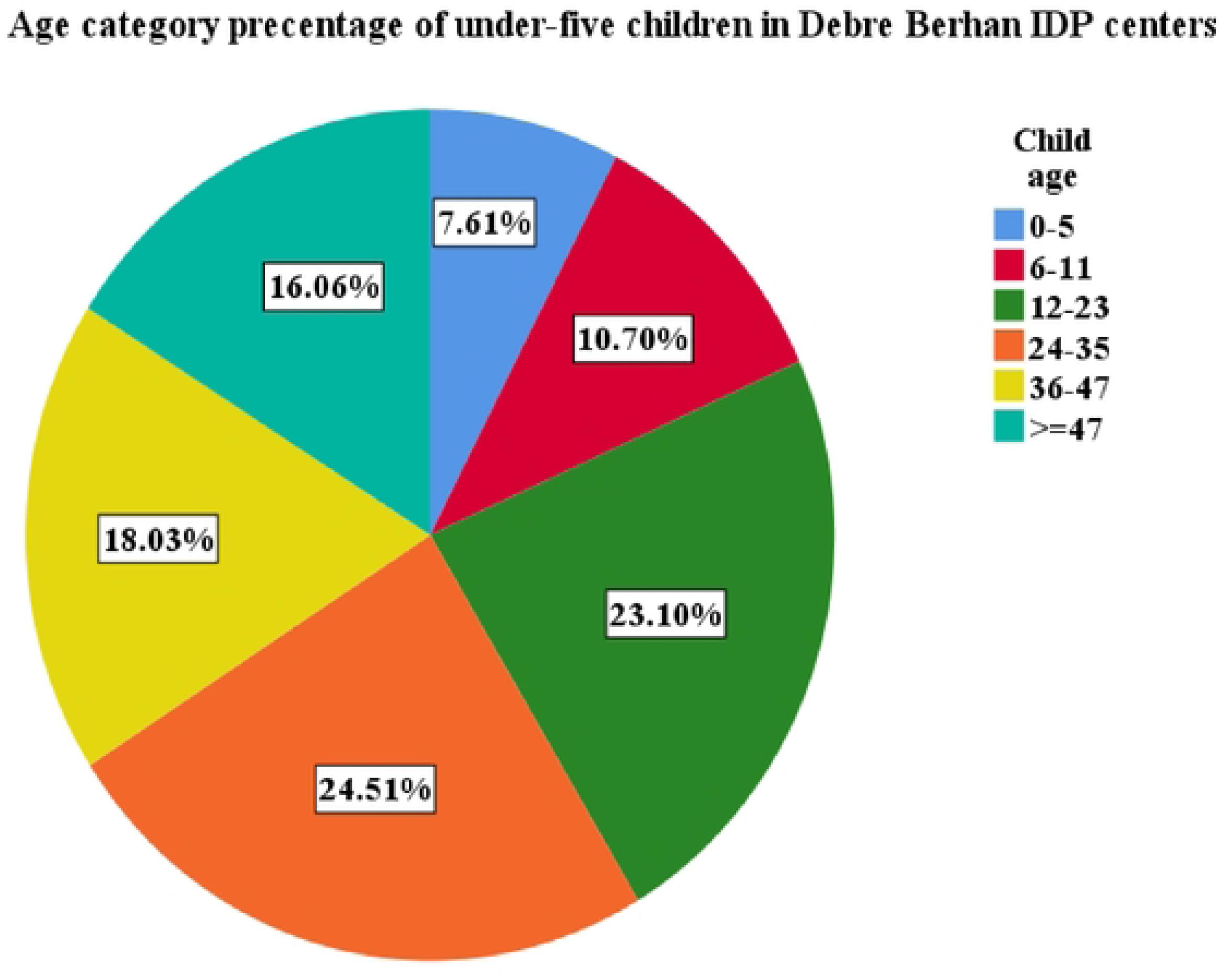
Age distribution of under-five children study participants in Debre Berhan IDP center, 2025.

### Behavioral factors

Handwashing practice shows that nearly three-fourths 262 (73.8%) wash after using the toilet. More than half 194 (54.6%) of them disposes their household wastes on open surrounding. About 283 (79.7%) had exclusive breastfeeding experience and only 107 (30.1%) breastfed according to recommendations. The majority 291 (82%) of them know at least one method of prevention of diarrhea. Among different method of prevention of diarrhea, more than three-forth 271 (76.3%) use handwashing. About 129 (36.3%) were partially vaccinated, and 24 (6.8%) of respondents were not vaccinated for the rotavirus vaccination (Table 2).

**Table 2:**
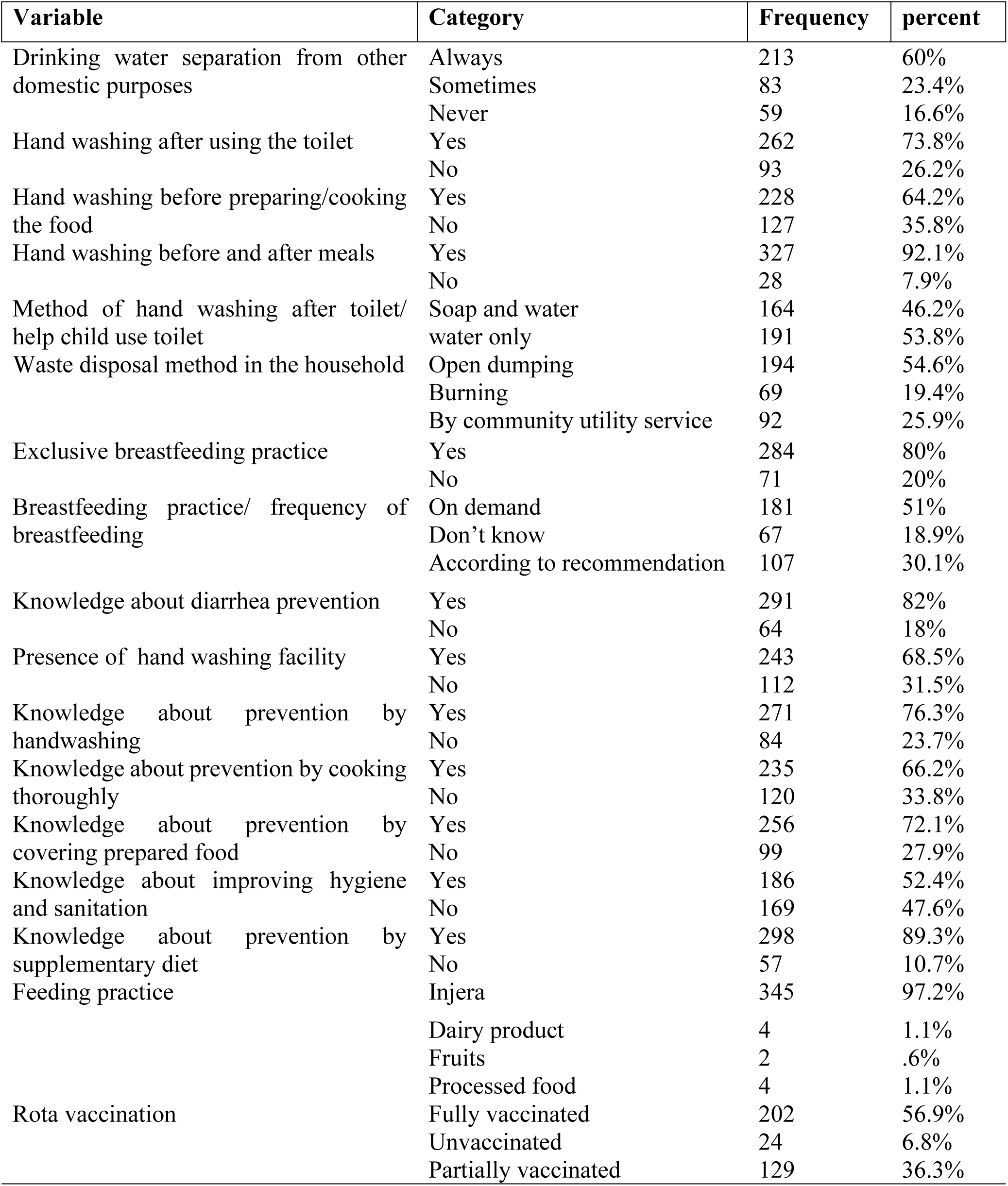
Behavioral characteristics of study participants in Debre Berhan IDP centers, (N= 355)

### Environmental factor

The majority 344 (96.9%) of them source of water was from public pipeline. However, storage methods rely almost exclusively on jerry cans 346 (97.5%). In addition, almost all of the respondents use latrine 349 (98.3%), and about 340 (95.8%) of them used shared/public type of latrines (Table 3).

**Table 3:**
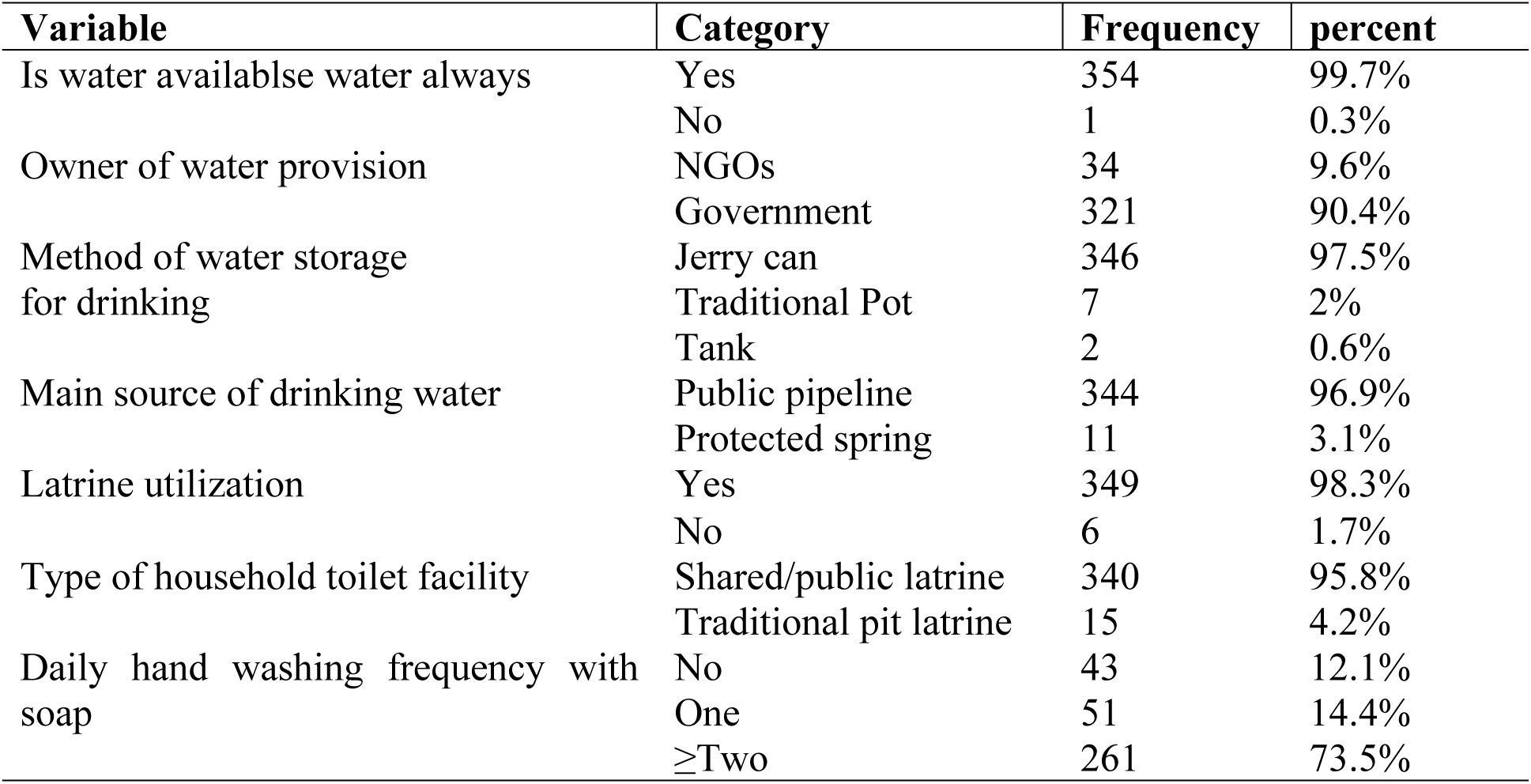
Environmental characteristics of under-five children in Debre Berhan IDP centers, (N= 355)

### Health facility and child care related factors

The coverage of key preventive and therapeutic health interventions was sub-optimal. Only 177 (49.9%) of children had received the recommended combined Vitamin A and deworming supplementation, and only one third 40 (34.8%) of them had received zinc supplementation. The vast majority of these cases 103 (89.6%) were lasting less than 14 days, while only 12 (10.4%) persisted for 14 days or more. Clinically, watery diarrhea was the most common presentation, accounting for 84 (73%) of cases, followed by bloody diarrhea 16 (14%) and mucus-containing diarrhea 15 (13%). Assessment of dehydration status revealed that about 47 (40.8%) exhibited some dehydration, and 4.4% were severely dehydrated, indicating a need for urgent medical intervention (Table 4).

**Table 4:**
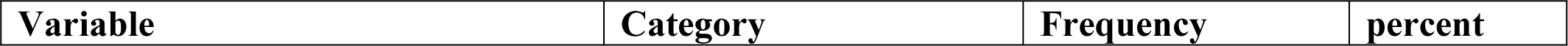
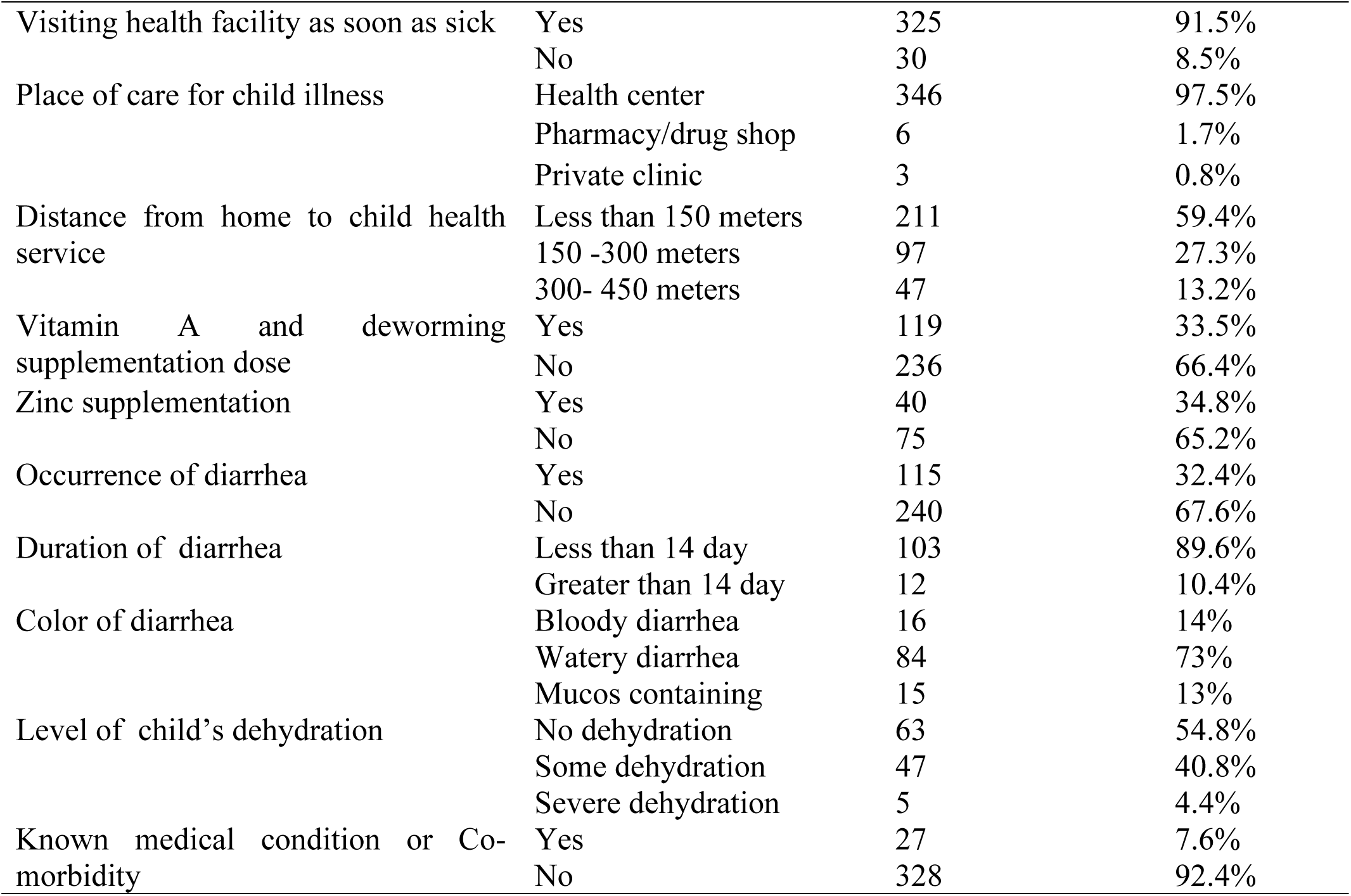
Health facility and child care-related characteristics of under-five children in Debre Berhan IDP center, (N= 355)

### Nutritional factors

Anthropometric assessments revealed 54 (15.2%) of children had global acute nutrition, with 17 (4.8%) severe acute malnutition and 37 (10.4%). MUAC assessment of children above six months also showed 20 (6.1%) and 34 (10,4%) had severe and moderate acute malnutrition, respectively. Fourteen (3.9%) children had severe undernutrition, and 29 (8.2%) had moderate underweight, whereas chronic malnutritional status shows 27 (7%) severe and 42 (11.8%) moderate stunting (Table 5).

**Table 5.**
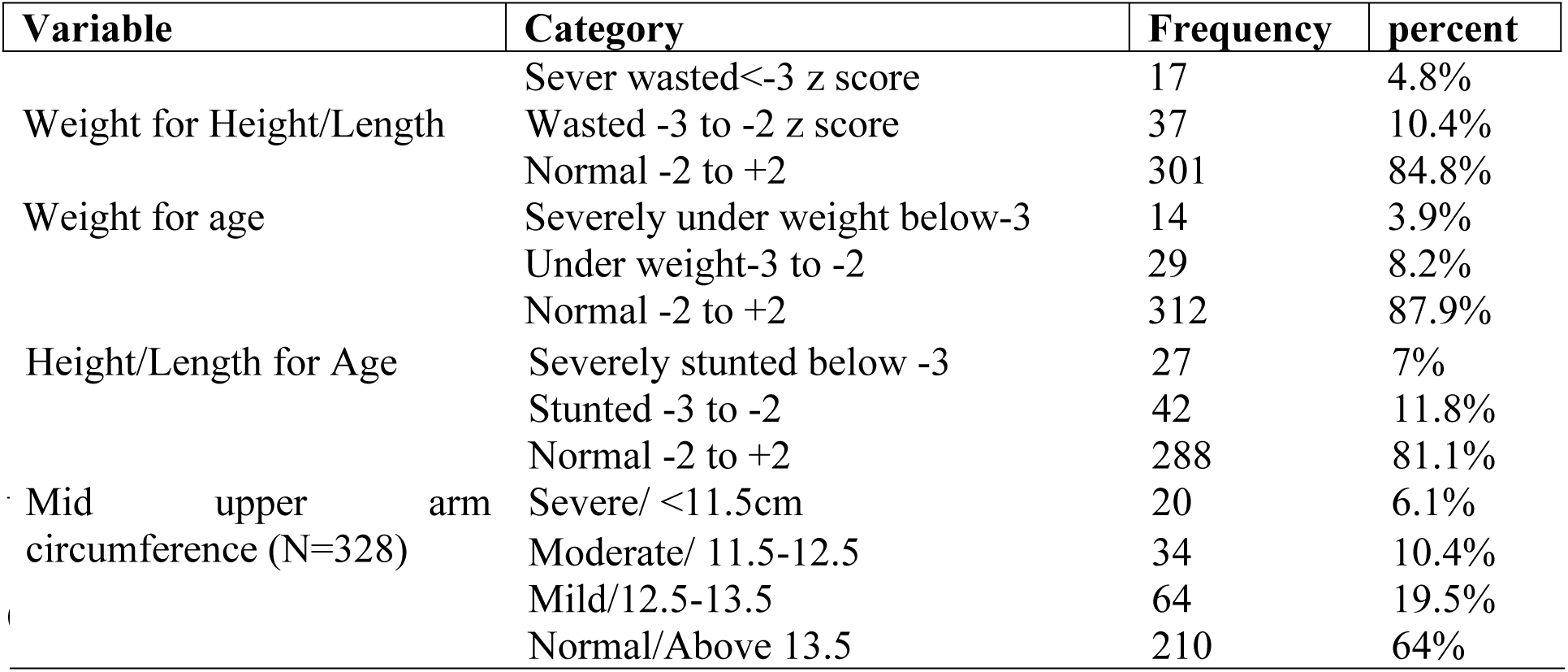
Nutritional characteristics of children under five years in Debre Berhan IDP center, (N= 355)

### Factors Statistically Associated with Diarrhea among Under-Five Children

The bivariable and multivariable analyses identified several key variables as significant independent predictors of diarrhea in Debre Berhan IDP center. In binary logistic regression, seventeen variables were showing association with diarrhea at p-value of <0.25. In Multivariable logistic regression, three variables, namely lower educational status of the mother or caregiver, waste disposal method in the household, and vaccination status were significantly associated with diarrhea at a p-value <0.05 (Table 6).

**Table 6.**
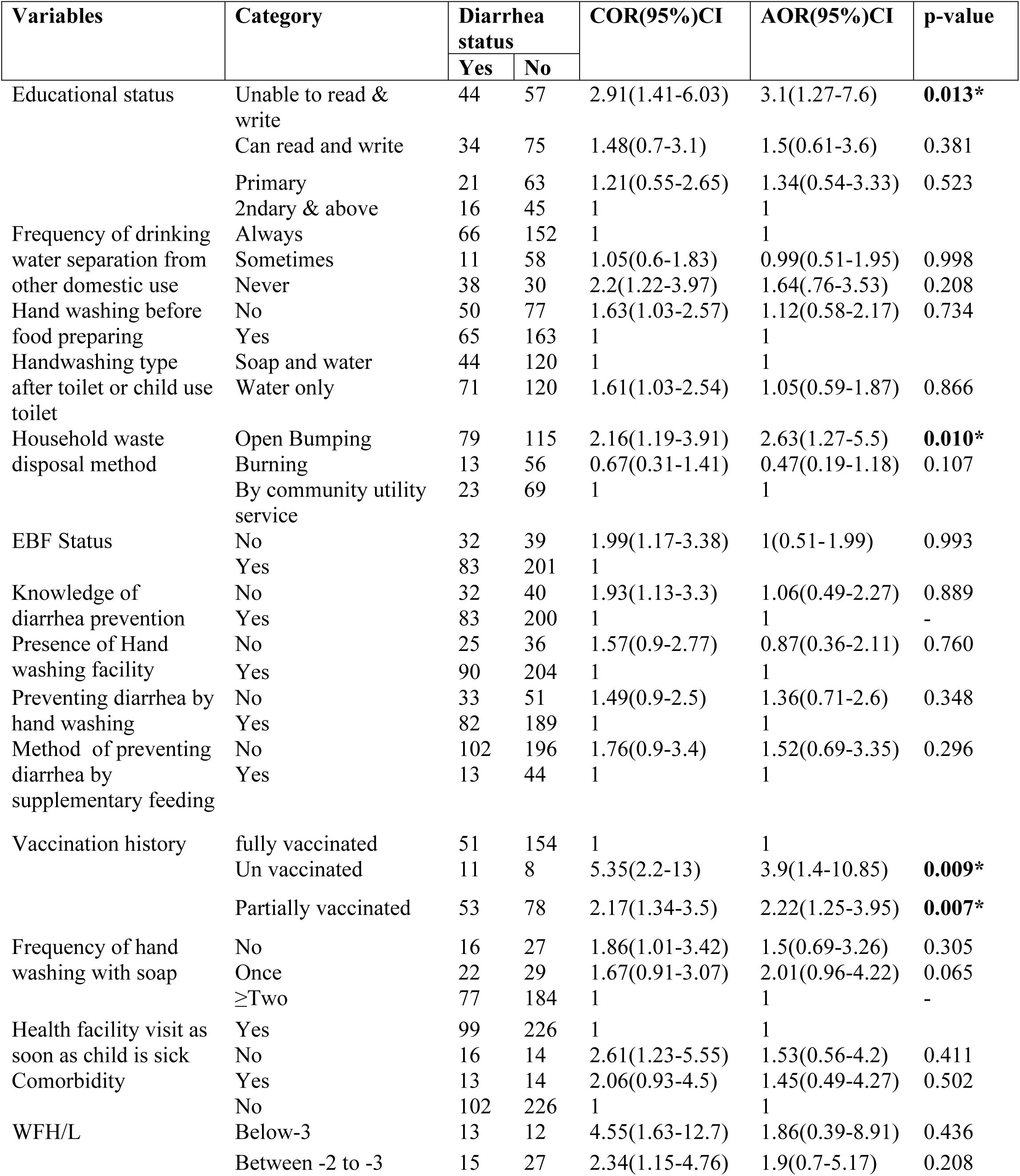
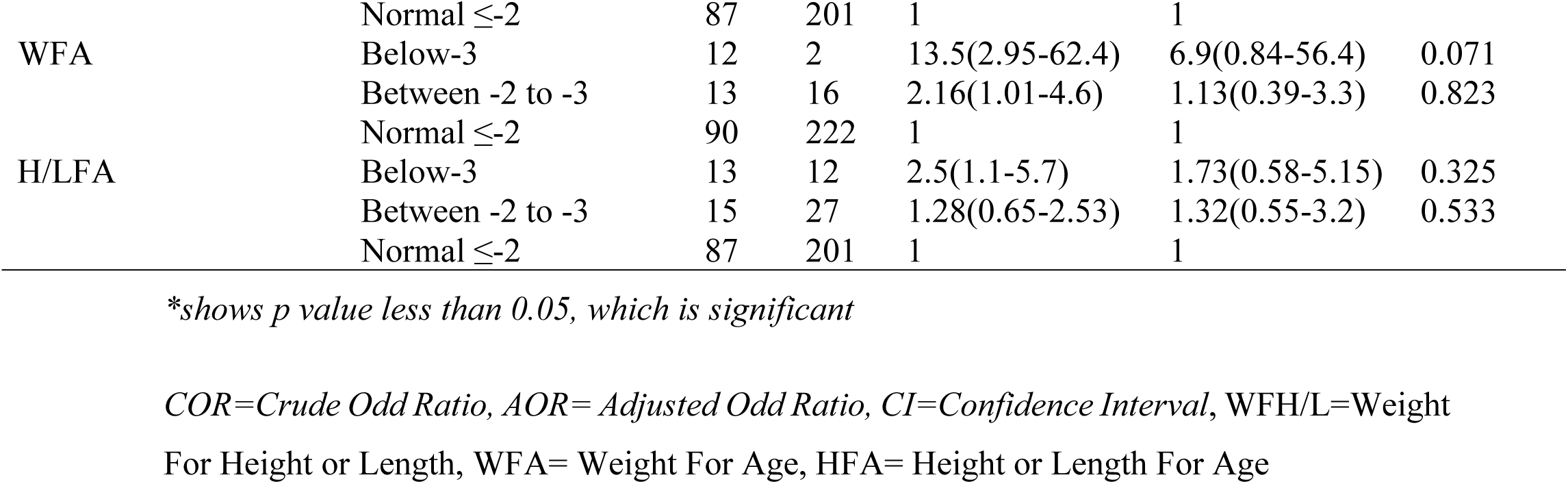
Bivariable and multivariable logistic regression analysis of factors of diarrhea among under five children at Debre Berhan IDP centers, (N=355)

Lower educational status of caregivers was associated with increased odds of diarrhea among under-five children. Mothers or caregivers who are unable to read and write were 2.91 times more likely to have diarrhea as compared to caregivers who had attended secondary school and above (AOR =3.1, 95%CI: 1.27-7.6, P=0.013).

Children from household practicing the open dumping waste disposal method had 2.63 times higher odds of developing diarrhea than households disposing their wastes by community utility service (AOR=2.63, 95%CI: 1.27-5.5, P=0.010).

Unvaccinated children had 3.9 times higher odds of developing diarrhea than fully vaccinated children (AOR=3.9, 95%CI: 1.4-10.85, P=0.009). Similarly, partially vaccinated children were 2.2 times more likely to develop diarrhea than fully vaccinated children (AOR= 2.22, 95%CI: 1.25-3.95, P=0.007).

The model fitness was checked with the Hosmer and Lemeshow goodness of fit test of the value 0.827, which is acceptable.

## Discussion

This study investigated the prevalence and associated factors of diarrhea among under-five children in Debre Birhan city Internally Displaced People’s centers. Thus, the prevalence of diarrhea among children under five in the Debre Berhan IDP center was 34.6%, which indicates diarrhea remains a major public health problem in IDP settings.

The finding of this study was nearly consistent with East Africa subgroup analysis 35% from a reviw report from IDP settings done in 2025 and the IDP camp Kahartum in Sudan (3,10). The possible similarity may be due to the fact that most IDPs in the region experience overcrowding, limited sanitation of water and hygine practice, inadequate sanitation facilities, malnutrition, lack of vaccination, and poor waste disposal practices. It can also be explained with shared socio-economic challenges such as poverty, low maternal educational status drought and instability in the region.

This finding was lower than the study done in Mekele IDP centers in North Ethiopia 52.3% (11), Gambella refuge camp in West Ethiopia 38% (18), Hargeisa IDPs center 51% in Somaliland (8), and unpublished report from Hodan district of Somalia 48%. The possible difference could be the availability of humanitarian aid, seasonal differences, study area, and access to water and sanitation services. However, the finding of the current study was higher than studies, such as a review report of 27% in IDP centers and 32% in refugee camps (3), 15.9% East Africa (19), 23.59% in Africa (9), and 22% and 19.62% in Ethiopia (6,12) The discrepancy may be due to poor waste management, lack of vaccination service like rota, limited access to safe drinking water, and overcrowding, which facilitates transmission of diarrheal disease in IDP centers. Similarly, our finding was higher than community based studies done in different part of Ethiopia ranging from 11-27.3%, (5,14–16,20–23) and Nigeria (24). The possible explanation may be, children living in IDP centers are more exposed to overcrowding, unsafe water source, inadequate sanitation facilities, and poor waste disposal. Furthermore, the difference might be due to sample size, study period, and cultural differences.

The multivariable binary logistic regression analysis identified three independent, associated factors that were significantly associated with the prevalence of diarrhea. Caregivers who are unable to read and write were 2.91 times more likely odds of having diarrhea than caregivers who had attended secondary school and above. This finding is in line with studies conducted in other countries, Kenya (25,26) India (27), and Ethiopian studies (11,14,16). Similarly, our finding was also supported by a systematic review and meta-analysis, which reported that children of mothers with no formal education were 2.5 times more likely to develop diarrhea as compared to children of mothers with formal education (12). This suggests that high diarrhea prevalence was associated with the educational status of the caregiver. This might be because an educated mothers may show a positive influence on safe drinking water, good sanitation, hygiene, and vaccination. In addition, the level of education helps caregivers to access and understand health care information and helps mothers to have better awareness of the manifestation of diarrhea, knowledge of preventing diarrhea, and seek early treatment for diarrheal disease.

Children from households practicing open dumping waste disposal method had 2.63 times higher odds of developing diarrhea than households disposing of their waste by community utility service. This finding was consistent with several prior community-based studies done in Ethiopia, (11,14,16,28–30), and low-resource settings from Nigeria (24), Kenya(25), Uganda (31), Bangladesh (32), and a Multi-country report (33). This might be due to improper waste disposal method of open dumping serves as a favorable condition for flies, rodents, and other disease vectors, which can contaminate food, water, and the environment by infectious agent causing diarrhea.

Unvaccinated children had 3.9 times higher odds of developing diarrhea than fully vaccinated children, and partially vaccinated children were 2.2 times more likely to develop diarrhea than fully vaccinated children. The report of this study was congruent with studies from Ethiopia (6,15,30,34,35), which reports that children who were not vaccinated or partially vaccinated for rota vaccine had a higher likelihood of developing diarrheal disease. The possible similarity across studies could be that IDP settings often have limited vaccination coverage, and inadequate vaccination increases susceptibility to key vaccine preventable disease such as rotavirus and measle virus, which causes diarrhea due to lack of protective immunity from immunization.

### Strength and Limitations of the Study

The cross-sectional study design can not show cause and effect relation ship. There might be a possibility of recall bias on diarrhea and vaccination history and social desirability bias on hygienic factors, which may underestimate or overestimate the finding. Since the study was conducted in winter, it doesn’t show the seasonal variation in the prevalence of diarrhea. Despite this, the study was the first study conducted in Amhara region, particularly in Debre Berhan town, which gives new insight into the extent of diarrhea among under-five children in IDP centers.

## Conclusion and Recommendations

The prevalence of diarrhea among children under-five years in the Debre Berhan IDP centers was 32.4% which is high. Maternal educational status, waste disposal practice, and vaccination status were independent factors found to be affecting the prevalence of diarrhea. Addressing these determinants through a comprehensive health education program, improving waste disposal practices and vaccination can contribute to a substantial reduction in diarrhea among children under five years. This finding suggests that integrated efforts are needed from concerned governmental and non-governmental organizations to enhance awareness by providing health education by focusing on improving water safety and sanitation, improving waste disposal practice, and increasing vaccination coverage in IDP centers. In addition, this finding will serve as a baseline for future researchers who need to conduct a longitudinal study to show causation by considering seasonal variation.

## Data Availability

Data will be available upon request from the corresponding author.

## Authors contribution

**Asaye Worku** has participated in the design, conception, tool preparation, data analysis, interpretation and in writing the manuscript. **Tsegaamlak Kumelachew** has participated in conceiving the study, perform data collection, data analysis, interpretation and revision of the manuscript. **Eleni Dagnaw, Tewodros Mulugeta, Abebe Nigussie and Hibist Kinfe** have designed the proposal, supervised the data collection, carried out the data analysis, investigate and interpreted of the result. All authors carefully read and approved the manuscript to be published in the journal.

## Availability of data and materials

Data will be available upon request from the corresponding author.

## Abbreviation

AOR: Adjusted Odds Ratio
COR: Crude Odds Ratio
CI: Confidence Interval
IDP: Internally Displaced People
SSA: Sub-Sahran Africa
SPSS: Statistical package for social science

## Acknowledgement

The authors would like to acknowledge Debre Berhan University, Asrat Woldeyes Health Science Campus for providing institutional support and ethical oversight necessary to conduct this research. We express our gratitude for Debre Berhan city administration health office for being cooperative in providing information regarding IDPs centers. The authors would also thank study participants for their willingness to participate in the study and data collectors for their commited work.

